# COVID19 epidemic modelling and the effect of public health interventions in India-SEIQHRF model

**DOI:** 10.1101/2020.09.16.20182915

**Authors:** Pooja Sengupta, Bhaswati Ganguli, Aditya Chatterjee, Sugata Sen Roy, Moumita Chatterjee

## Abstract

A dynamic epidemic modeling, based on real time data, of COVID19 has been attempted for India and few selected Indian states. Various scenarios of intervention strategies to contain the spread of the disease are explored.

## 1 Introduction

Corona Virus Disease 2019 (COVID-19) caused by the novel corona virus SARS-CoV-2 has been found to be catastrophic leading to loss of life along with sacrifice of major economic activities throughout the world, from developed to developing nations and from poor to rich nations. It has been declared by World Health Organization a pandemic [Mission [2020]]. The disease, originated at Wuhan in the province Hubei of China and has rapidly spread first in selected parts of eastern Asia including South Korea and Japan and then to entire Asia, Europe and USA and other parts of the world. Its global reach and devastation are unprecedented and continue to accelerate unabated leading to loss of life and global suspension of entire economic and social activities. This is considered to be the worst kind of disaster for humanity in recent times. It’s expected zoonotic origin coupled with lack of vaccine and treatment barring some palliative care have made its management extremely difficult. Till date nearly 2.34 million people have been affected, with the number of deaths being nearly 0.16 million, leading to a gross case fatality rate of 6.89 % among the infected who have been diagnosed with the disease.. According to https://www.worldometers.info/coronavirus/ on third week of April, 2020, the prevalence rate of the disease is 299 per million population, while the prevalent death rate of the disease is 20.6 per million population. Among the active cases of 1.58 million, 4% are critical while among the closed cases of 0.76 million, 21% death has been reported. In India, by that time, in all 16365 cases have been reported with 521 deaths, while the first case has been detected on 30th January, 2020.

Owing to this extremely difficult situation towards management of the disease, the administration and the medical community have recommended lockdown and social distancing at various stages and phases with a combination of home confinement of population, suspension of all natural human activities and movements barring a few emergencies along with sanitization as possible ways of containment and mitigation of the spread of the disease. The disease curve is obtained by plotting the prevalence of the disease over time which is expected to reach a peak, become stable and then decrease monotonically. Any mitigation measure is aimed towards putting a threshold in the magnitude of the peak and delay in attaining the same, followed by stability which is technically known as ‘flattening of the curve’. To prevent the stage III or community spreading of the disease, almost all countries have imposed variable number of lockdown days that ranges from 4 to 6 weeks to achieve such flattening. Depending on the real-time scenarios, experts are in the process of updating the lockdown period. As an example, in India the lock-down duration first clamped from March 24 to April 14, 2020 and later extended till May 3, 2020. However, deciding on the optimal or ideal number of lockdown days is an extremely difficult task as both sides the life and loss of economic activities are to be considered. Owing to the novelty of the virus, the human civilization has no prior knowledge to gauge the consequences. However, based on earlier experiences to deal with other pandemics, viz SARS or MERS, the medical community has tried to gather understanding about them. Several research groups are working on the evaluation of such containment measures by predicting the course of COVID-19 cases in India, under various scenarios (Ref: Chatterjee et al [2020], Das et al [2020], Das [2020], Ghosh et al [2020]). In the present work we have attempted such scenario profiling through extensive simulation experiment for the assessment of the mitigations and their consequences in India and a few selected states, where the disease prevalence is high.

## 2 Model

Susceptible-Infectious-Recovered (SIR) models are popular in predicting the course of epidemics (Ref: Smith Moore [2004], Allen et al [2008], Korobeinikov [2009]), where the population is divided into 3 compartments of Susceptible (S), Infectious (I) and Removal (R) (through recovery or death) groups with a defined rates of transition among them. Variation of SIR model is the SEIR model which includes another compartment as Exposed (E) population in the model (Ref: Stehlé et al [2011], Li et al [1999], Röst [2008]). Recently an extension of the SEIR model has been proposed (Ref: Churches [2020], Ghosh et al [2020]), where apart from the above mentioned 4 stages, two other stages viz. Quarantine (Q) and Hospitalization (H) are considered to take in account the healthcare capability and the R stage has been segregated to usual Recovery (denoted by R) and fatality (denoted by F). The description of individual compartments are as follows.

| State | Indicator | Functional Definition |
| --- | --- | --- |
| Susceptible | S | Susceptible individuals to COVID-19 |
| Exposed | E | Exposed and infected, not yet symptomatic but potentially infectious |
| Infected | I | Detected with COVID-19 who are infected symptomatic and infectious |
| Quarantined | Q | Infectious and self-isolated (individuals who are isolated and hence do not come in contact with the susceptible population) |
| Hospitalized | H | Requiring hospitalization (would normally be hospitalized if capacity is available) |
| Recovered | R | Recovered, assumed to be immune from further infection. However, repeat infection is possible, but chances are low and for the time being assumed ignorable. |
| Fatality | F | Case fatality (death exclusively due to COVID-19 and of no other cause. |

In the present study we will fit the above mentioned compartment model to COVID 19 outbreak data for India and use simulations to explore the impact of lockdown on the epidemic and perform a what -if analysis for strategies for imposition and relaxation of the lockdown.

### 2.1 Choice of parameters

The resulting curves are validated with the observed data for India from https://ourworldindata.org/coronavirus-data for the pre lockdown period and the best fit is obtained to the observed case and fatality data by using the following set of parameters,

- Number of exposure events per day between symptomatic infected and susceptible = 2 (low value as India started implementing screening and quarantine for suspected cases from mid January)
- Probability of passing an infection from infectious to susceptible =0.03
- Number of exposure events per day between asymptomatic and susceptible = 40 (relatively higher value as the testing rate was low)
- Probability of passing an infection from asymptomatic to susceptible =0.01
- Number of exposure events per day between quarantined and susceptible = 2
- Probability of passing an infection from quarantined to susceptible = 0.02
- Rate per day at which symptomatic infected enter quarantine =1/5. (as the Indian government was both quarantining and advocating self quarantine from January)
- Available hospital beds for the susceptible whom each case infects = 40
- Daily death rate among susceptible = 7.3/1000/365 (based on CDR for India)
- Daily death rate among asymptomatic = 20/1000/365 (as they are probably in early stages of infection)
- Daily death rate among quarantined = 30/1000/365
- Daily death rate among infected = 30/100/365 (As deaths will occur in the H compartment. Note also that H denotes needing to be hospitalized and not necessarily hospitalised as all may not seek care or have access)
- Daily death rate among hospitalised = 80/365/1000.

Note that the model also includes two additional compartments corresponding to asymptomatic individuals and cases requiring hospitalisation which would put additional burden on the healthcare system. The special interest for this model is the E compartment of asymptomatic infectious whose behaviour over time can account for increased case numbers even after improvement in healthcare facilities for symptomatic cases.

## 3 All India analysis

Figure 1 present the epidemic curves from the fit for the pre-lockdown period in India. Time is indexed as starting from the date of the first confirmed case corresponding to January 30th, 2020. The fitting has been done based on the extensions to the EpiModel package in R developed by Tim Churches (https://timchurches.github.io/blog/posts/). As initial confirmed cases largely corresponded to foreign travellers, each such case is used as the index case for infecting a network and the cumulative effect of the resulting transmission is presented below. Case specific data for India is obtained from www.coviv19india.org.

**Figure 1.**
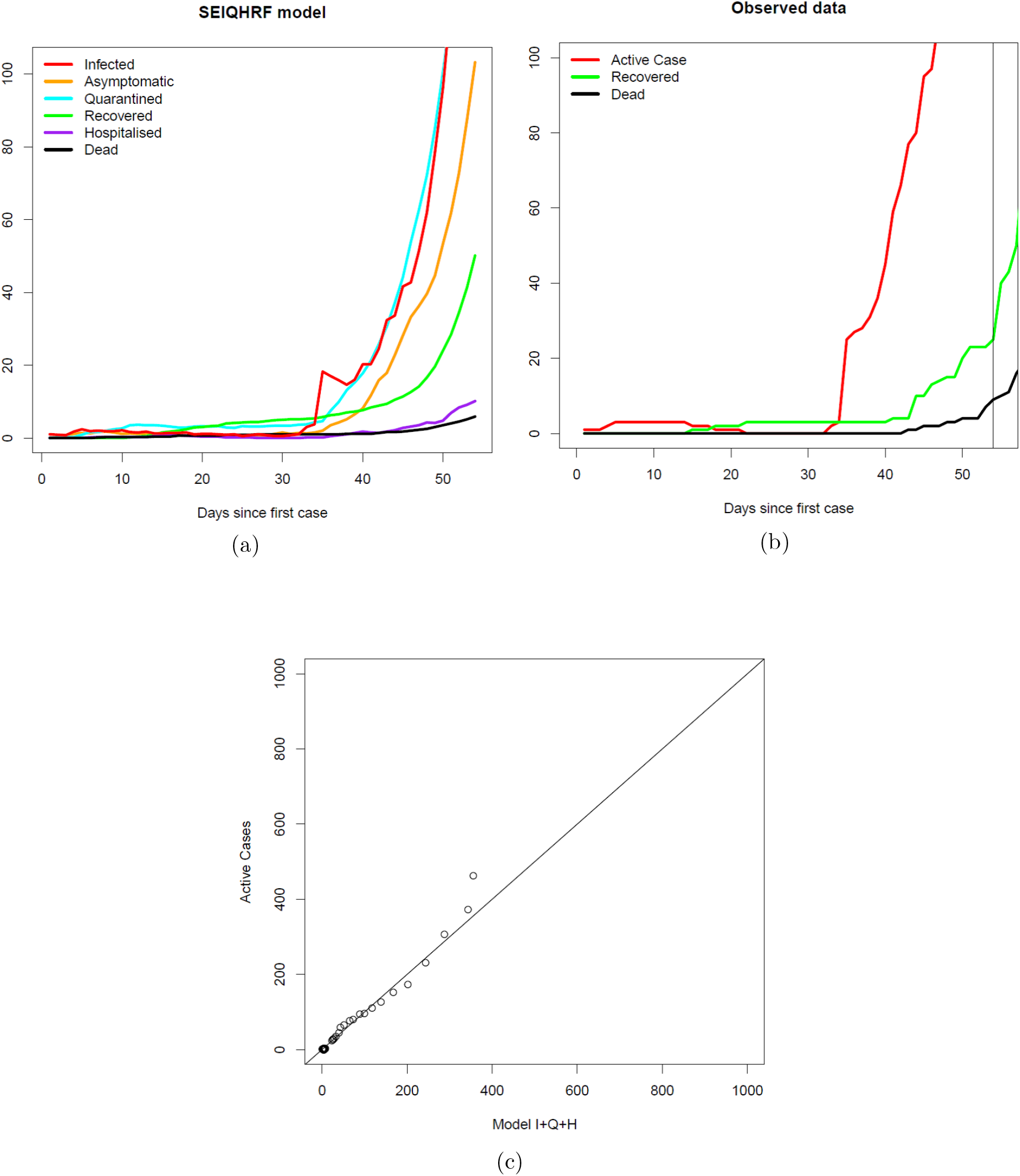

The above figure shows the predicted daily prevalence for the various compartments for the model, the plot of active, deceased and recovered cases for India pre lockdown and the agreement of the observed and model based daily case numbers. Note that the appropriate comparison is between the daily count of active cases and the sum of cases in the infected, quarantined and hospitalised compartments.

### 3.1 What if there was no lockdown and/or travel ban?

We shall next simulate from the fitted model to estimate likely case counts in the absence of a lockdown and a travel ban. We shall assume that infected travellers continue to enter India at the observed average rate over the pre lockdown period and observe the results until the first week of April.

The above figure shows the simulated case counts for the various compartments until the first week of April and a comparison of the model based predicted daily cases and the observed daily active counts. Note that the points in the second plot lie below the 45 degree line indicating that the active cases post lockdown are generally lower. This gives some preliminary evidence of the effectiveness of lockdown and travel bans. Note also that the model output suggests an enormous spike in asymptomatic cases which are potentially infectious. This suggests that one reason for the effectiveness of lockdown is reduction in the contact between such individuals and the susceptible population. If such a situation is to prevail until the end of May, the model estimates a total active case count of 45000 including 1300 fatalities.

**Figure 2.**
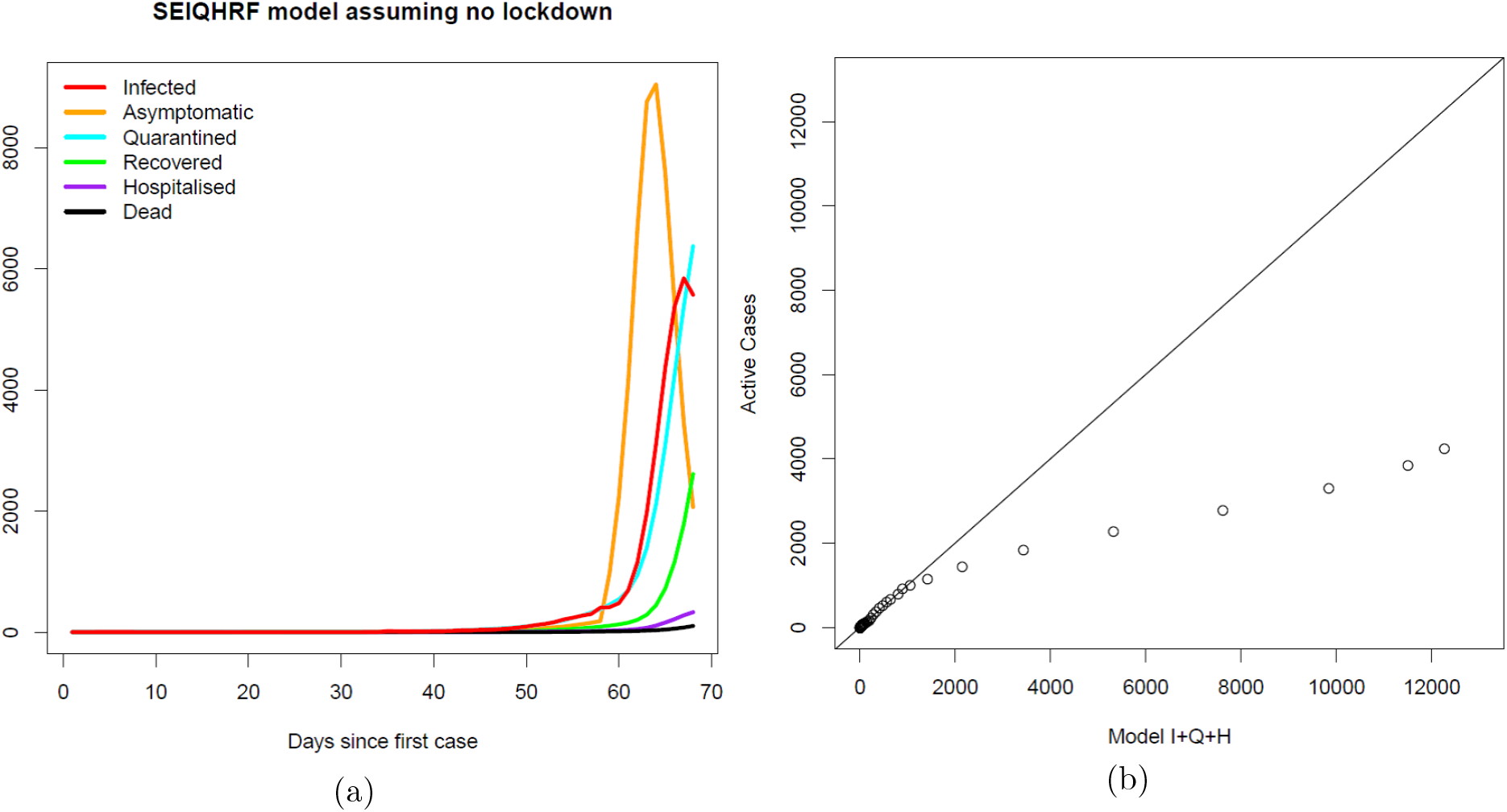

### 3.2 Did the early intervention strategy work?

The above simulation does, however assume that India’s early intervention of screening, contact tracing and quarantine is maintained throughout the period. Case numbers and fatalities have been far lower in India than in other nations of comparable population and the controversy regarding testing notwithstanding, this could be a potential alternative explanation for the low counts. We next explore the predictions for India assuming that the government had done nothing. The contact rate parameters between the various model compartments and the quarantine rate are adjusted to reflect this. The infection parameter is increased slightly to reflect the absence of public health information campaigns such as those promoting hand washing. Had the government adopted no strategy to combat the epidemic, the model estimates that at the end of May, the model estimates that total active case count could be as much as 1076881 including 113151 fatalities.

### 3.3 What should we expect if lockdown is completely lifted?

We refitted the best fit SEIQHRF model to the case data using the case counts upto the first week of April. The effect of lockdown is incorporated into the model by reducing the contact rate parameters between the various compartments. Relaxation of lockdown is modelled by restoring these parameters to their original values in the pre lockdown period. The earlier strategy of airport screening, contact tracing and quarantine is however assumed to continue. For this situation, the total case count is observed to be 85000 with 1500 fatalities at the end of May.

### 3.4 Staggered withdrawal of lockdown

To understand the impact of a staggered withdrawal of lockdown, a few more options are explored and a forecast of total case count and fatalities are obtained.

- Weekwise staggered relaxation of lockdown between April 14th-30th: For this situation, the total case count is observed to be 41560 with 920 fatalities at the end of May.
- Weekwise staggered relaxation of lockdown between April 14th-31st May: For this situation, the total case count is observed to be 33250 with 800 fatalities at the end of May.
- Extension of total lockdown by further 2 weeks after April 14th followed by complete relaxation: For this situation, the total case count is observed to be 26560 with 620 fatalities at the end of May.

## 4 Analysis of selected Indian states

After the all India profiling, the study has been carried out for 28 Indian states. The *act* parameter for each state has been adjusted based on the population density of the states. These 28 states have been ranked in decreasing order of *act*, thus one with higher value of *act* has more number of exposure events between infectious individuals and susceptible individuals.

In the baseline model, we see the result of our simulation of our hypothetical world of 1000 people. Our observations are;

- The epidemic subsides in about two months, provided the required assumptions hold.
- The population density of Delhi being very high, very high number of people are infected, although asymptomatic.
- Prevalence tends to start with exponential growth then tapers off.
- The number requiring hospitalisation seems reasonable, not too large.
- The number in the case fatality compartment is monotonically increasing, as expected, but at a much lower rate.

**Figure 3:**
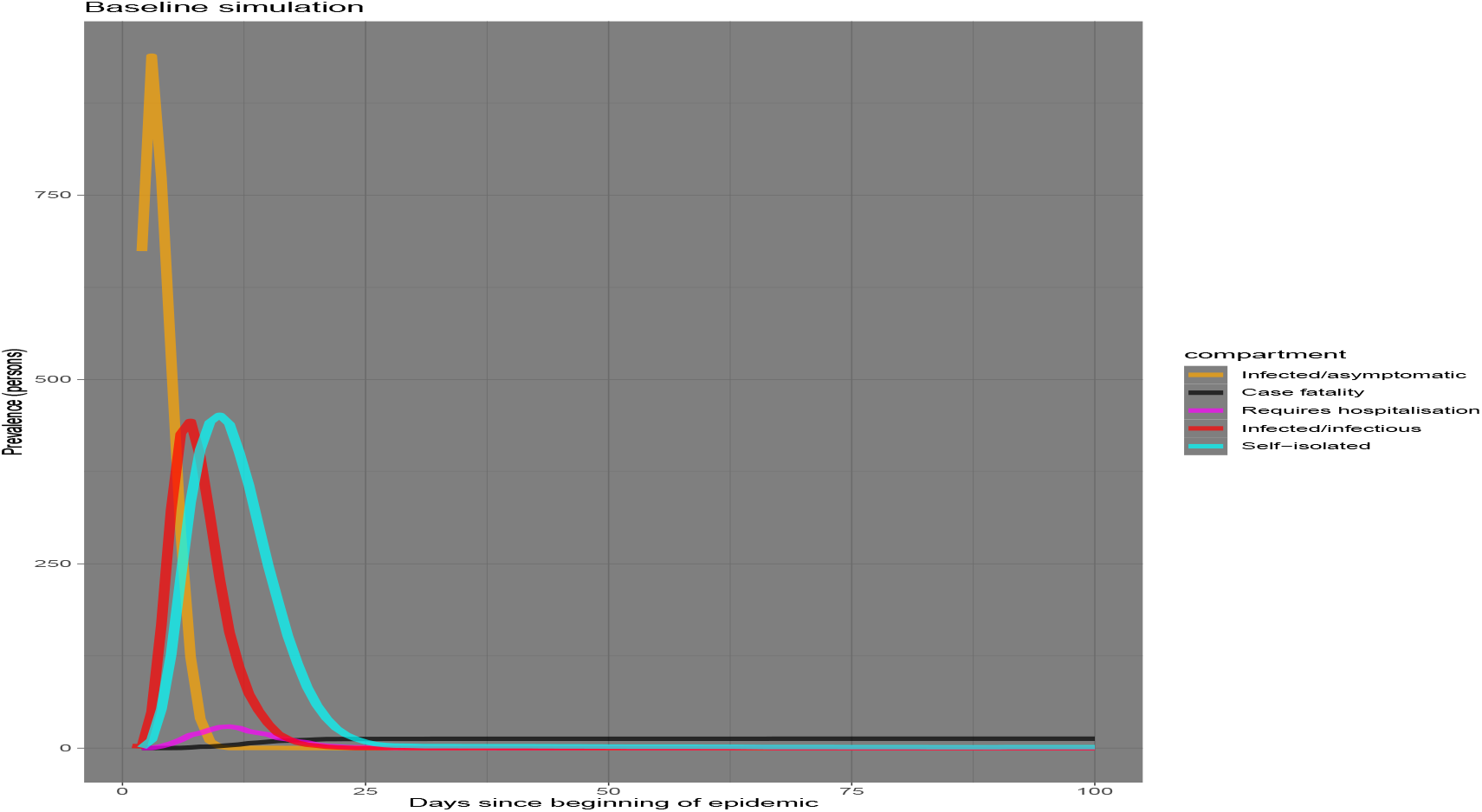
Prevalence of COVID-19 in Delhi population.

We then run the simulation under four different experimental set-up;

- Ramp up hospital capacity to triple the baseline level, starting at day 15.
- Step up social distancing (decrease exposure opportunities), starting at day 15, for everyone except the self-isolated, who are already practising it.
- More social distancing but starting at day 30.
- Increase both social distancing and self-isolation rate starting at day 15.

A comparative study of the four experimental intervetions with the baseline is shown in figure 5 for Delhi. Some obervations are;

- When we ramp up hospital capacity to triple the baseline level, starting at day 15, not much is seen as improvement in the number of infectious and asymptomatic. In fact not much change is seen in any of the compartments if only the hospital capacity is increased.
- With a step up in social distancing (decrease exposure opportunities), starting at day 15, for everyone except the self-isolated, shows a marked decrease in the number of infected/ infectious individuals.
- When, by day 30, social distancing is increased to halve the number of exposure events between the infected and the susceptible per day, there is a further reduction in the number of infected people. Also more number of people are self-isolated. Thus reducing the chance of infection further
- A combination of self-isolation and social distancing seem to be the most effective way of containing the spread of the disease.

**Figure 4:**
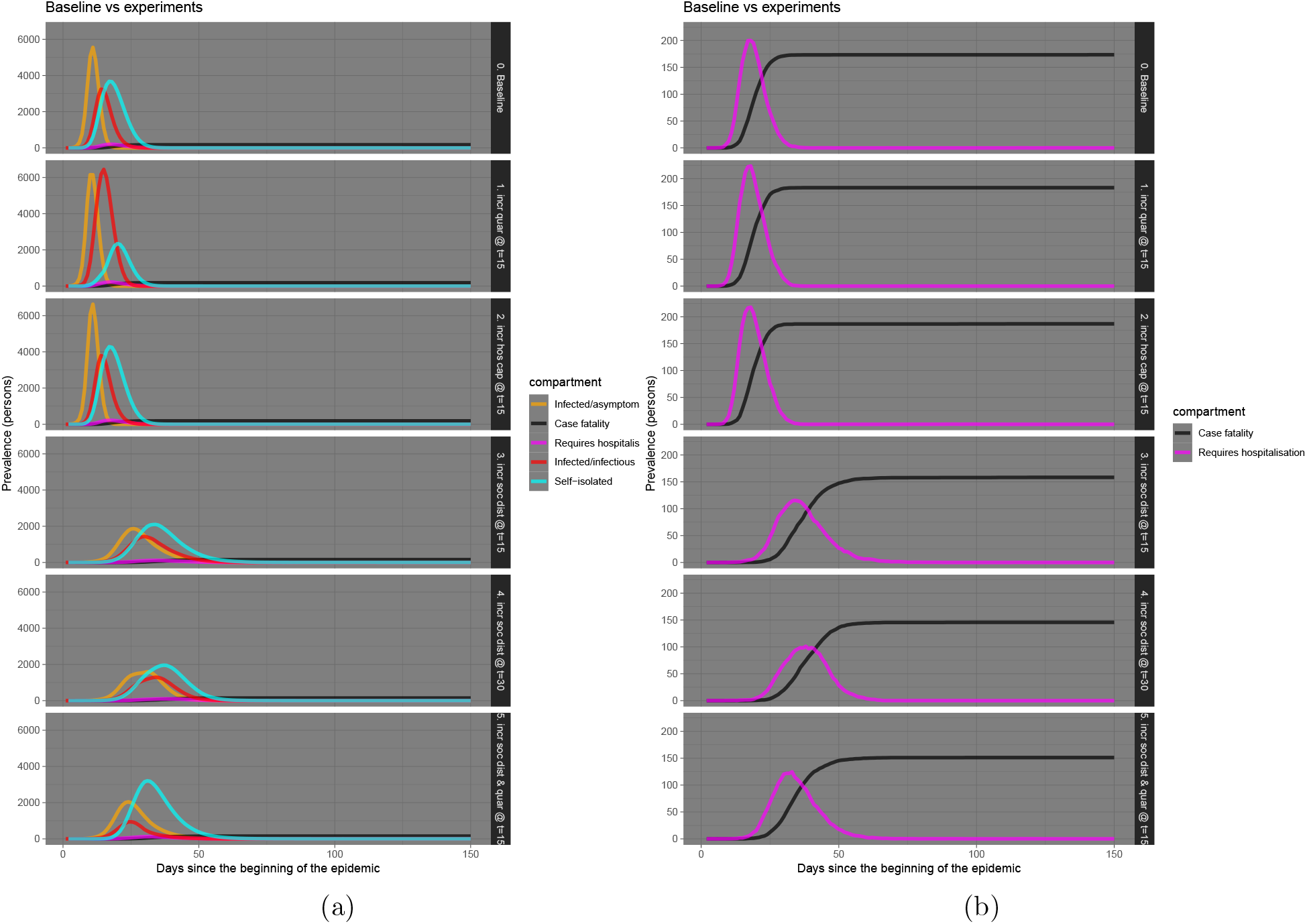
Delhi

**Figure 5:**
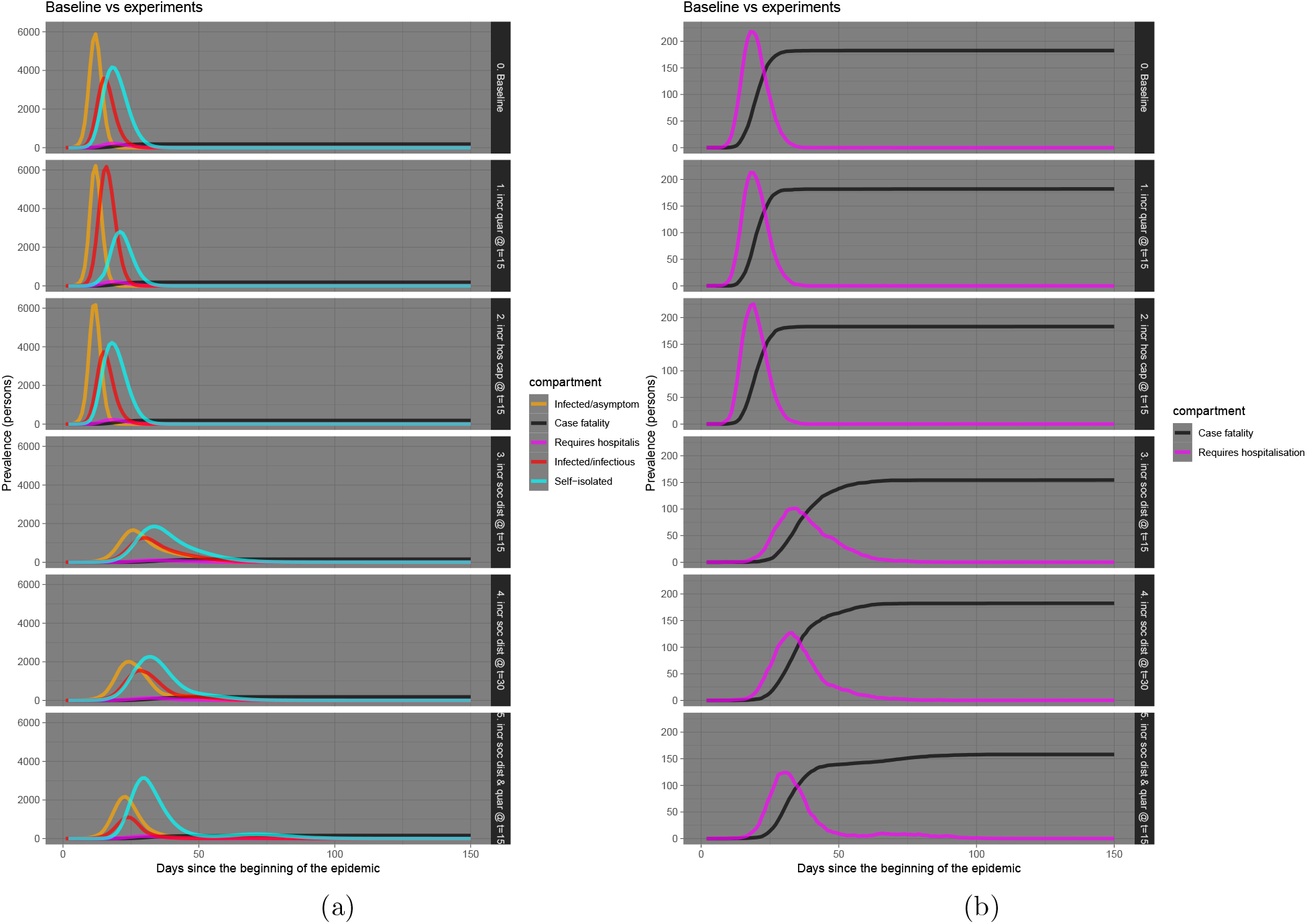
Kerala

The results from some selected Indian states are presented in figures 6 to 10. These states have been selected based on their population density; the figures represent states in decreasing order of their density. States with denser population have increased risk of exposure of susceptible individuals to infectious ones. The plots represent the difference in the growth rate of infectious, hospitalised and fatal cases. Of the selected states, Kerala has been to some extent successful in controlling the number of active cases and fatalities.

**Figure 6:**
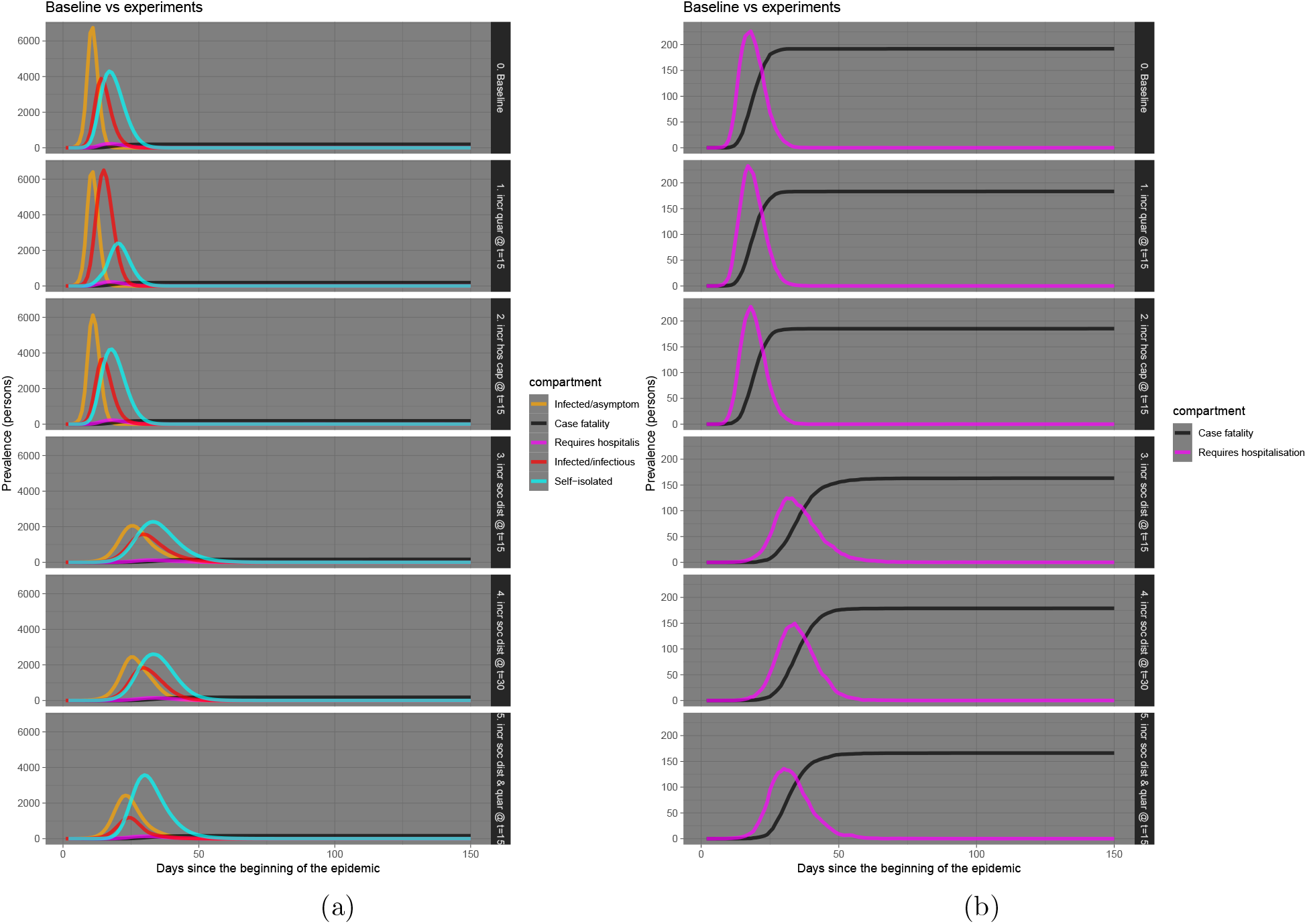
West Bengal

**Figure 7:**
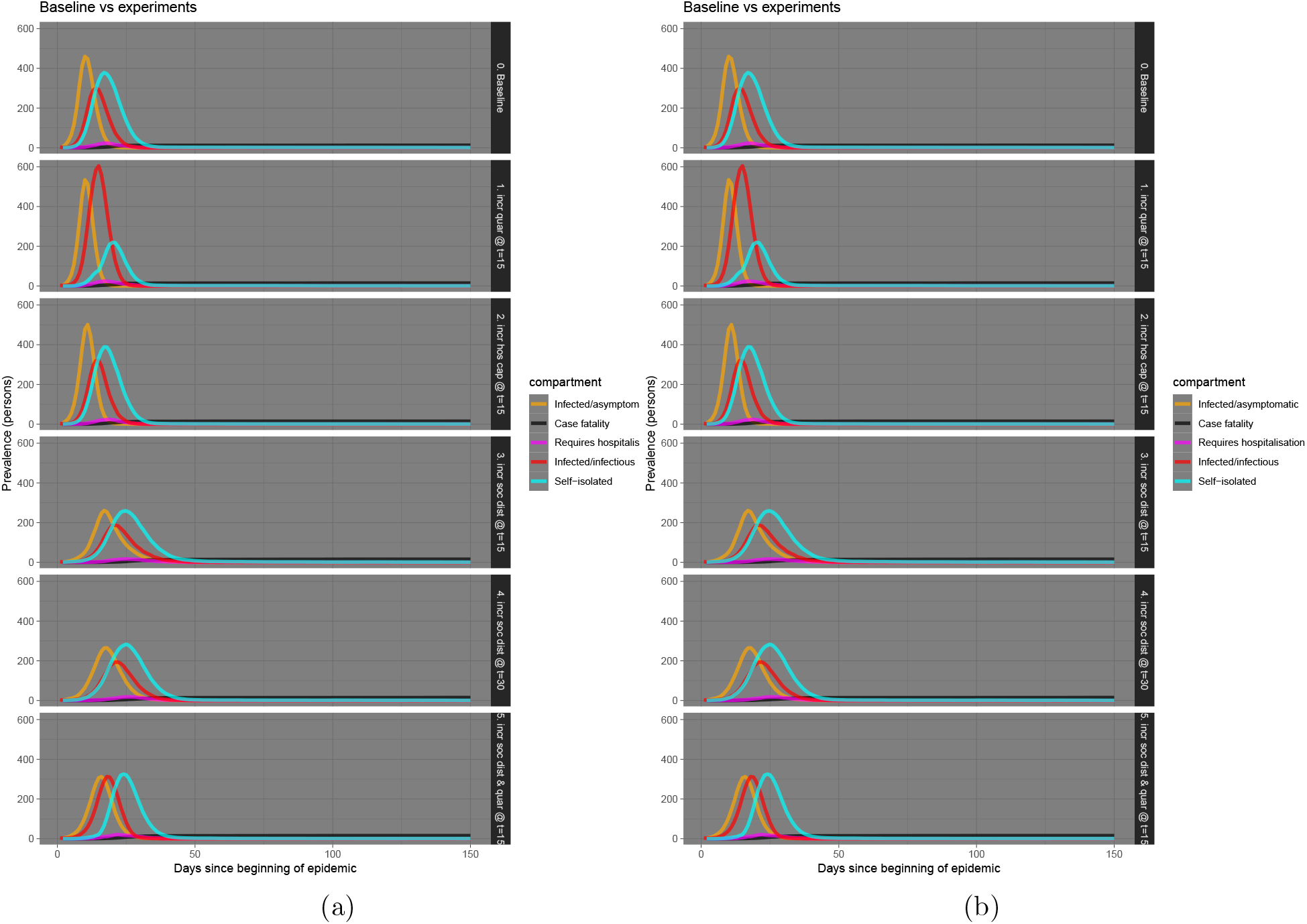
Haryana

**Figure 8:**
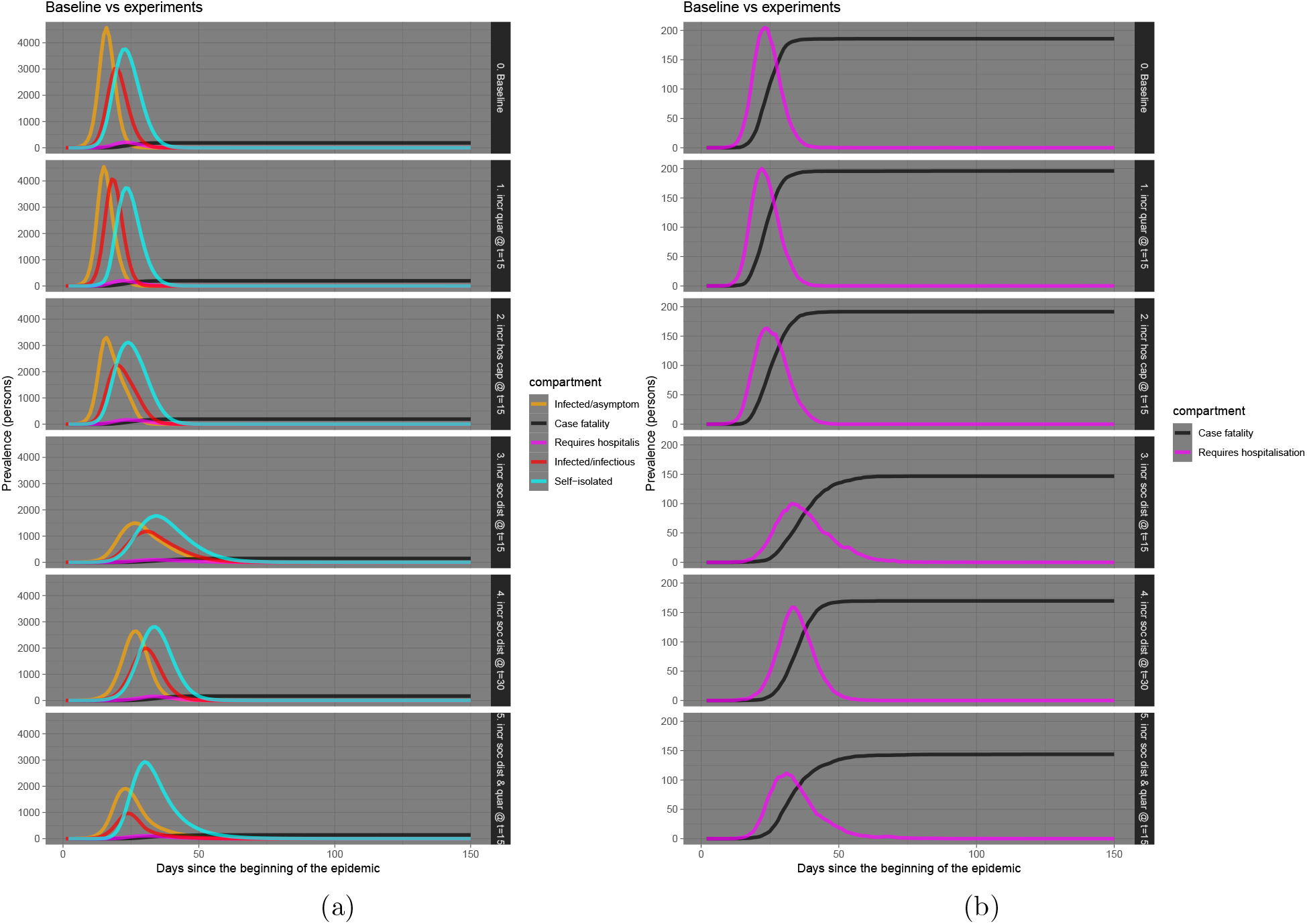
Tamil Nadu

**Figure 9:**
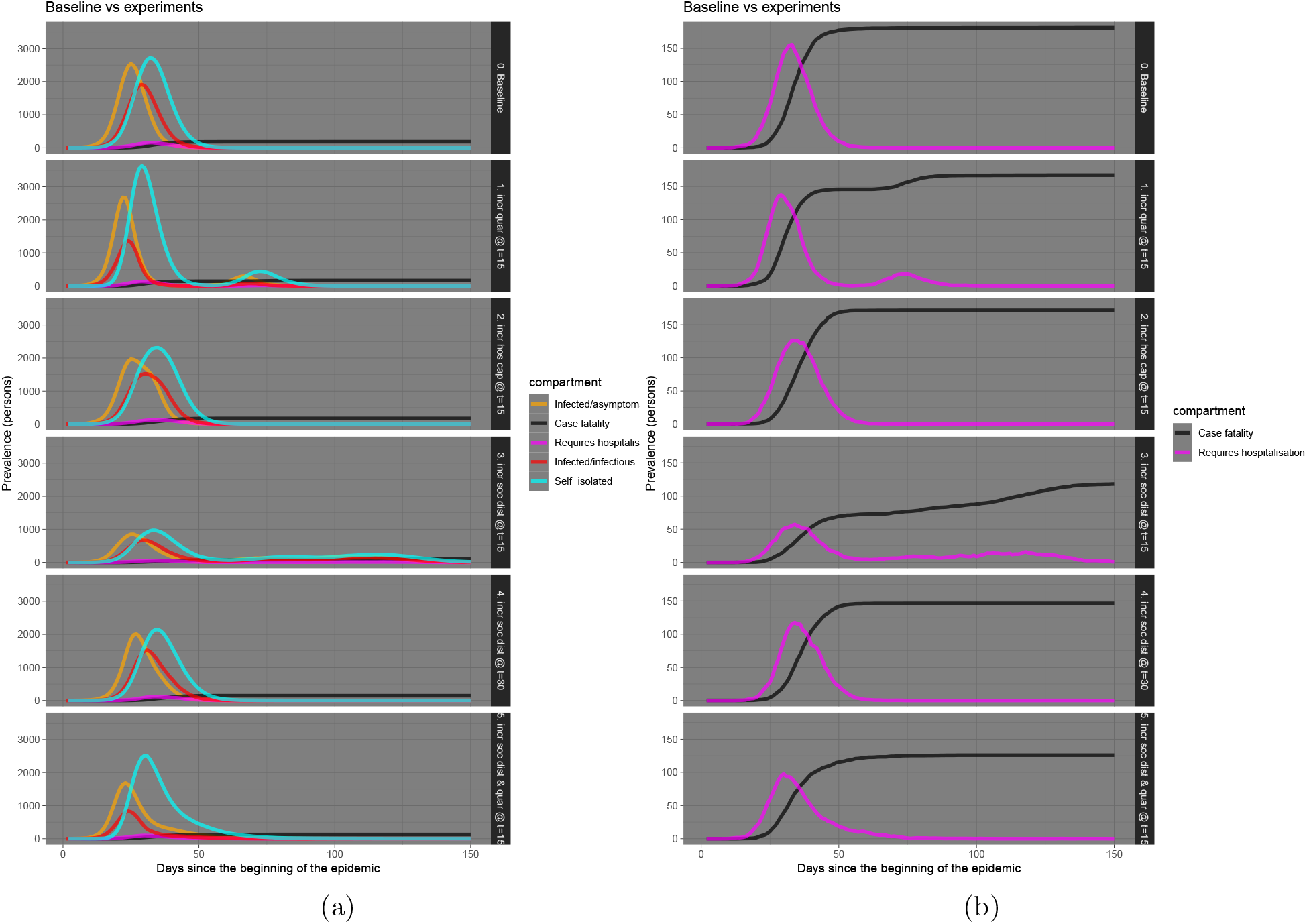
Telengana

**Figure 10:**
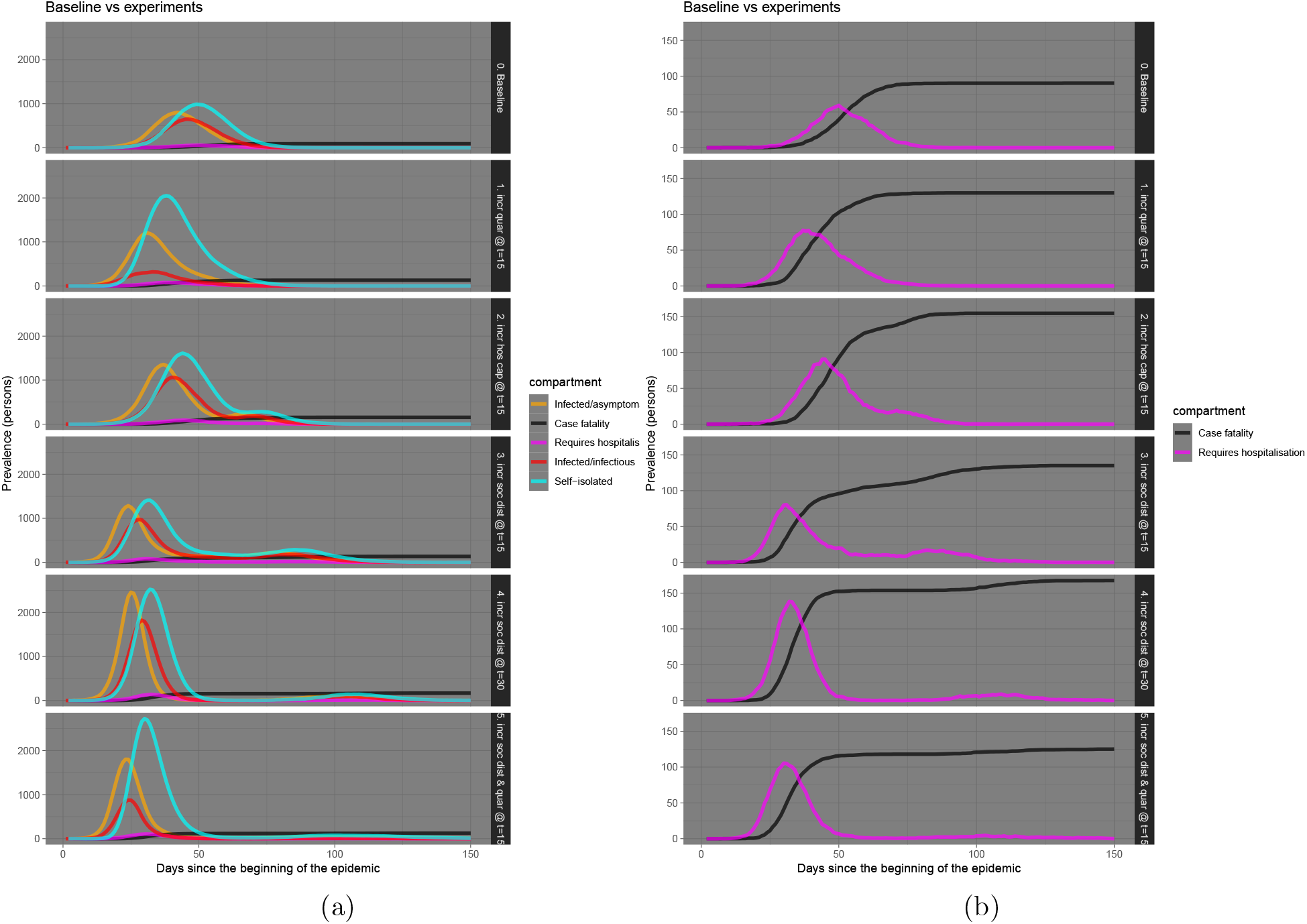
Rajasthan

## 5 Conclusion

In the present work we have tried to assess the Indian scenario of COVID19 outbreak with a refined dynamic model accommodating hospitalization and quarantine apart from the usual SEIR components. The scenario analyses of some of the important states are also performed. It has been projected that around two month span is required to arrest the epidemic with the present strategy of lock-down and social distancing that includes phased withdrawal of such measures depending upon the identified hot spots. A more detailed analysis with various other parameter choices is warranted to understand the ever changing nature of the epidemic. The present work is dependent on the best available parameter choices and only estimates of various entities are given. It is imperative that economic consequences of lock-down should be taken into account to suggest any policy decision, which is beyond the scope of the present work.

## Data Availability

The data used in this study are simulated for the necessary analysis.

